# Effects of recurrence of illness and adverse childhood experiences on effector, cytotoxic, and regulatory T cells, and cannabinoid receptor-bearing B cells in major depression, an autoimmune disorder

**DOI:** 10.1101/2023.06.11.23291243

**Authors:** Michael Maes, Muanpetch Rachayon, Ketsupar Jirakran, Atapol Sughondhabirom, Pimpayao Sodsai

## Abstract

**Background:** Major depressive disorder (MDD) is characterized by increased T helper (Th)1 polarization, T cell activation (e.g., CD71+ and CD40L+), and cannabinoid receptor type 2 bearing CD20+ B cells; and lower T regulatory (Treg) numbers.

**Aims:** To delineate the effects of adverse childhood experiences (ACEs) and recurrence of illness (ROI) on activated T and CB2-bearing B populations, and Tregs, including FoxP3+CD152+, FoxP3+GARP+, and FoxP3+CB1+ cells.

**Methods:** We measured ROI, ACEs, the number of activated T cells, Tregs, and CD20+CB2+ B cells, in 30 MDD patients and 20 healthy controls.

**Results:** A larger part of the variance in the depression phenome (40.8%) was explained by increased CD20+CB2+ and activated T cells, and lowered Tregs. ROI and lifetime suicidal behaviors were significantly and positively associated with CD20+CB2+, CD3+CD71+, CD3+CD40L+, CD4+CD71+, CD4+CD40L+, and CD4HLADR+ numbers. ROI was significantly correlated with CD8+CD40L+ numbers. The sum of ACEs was significantly associated with CD20+CB2+, CD3+CD40L+, CD4+40L+ numbers, T cell activation (positively) and Treg (inversely) indices. One replicable latent vector could be extracted from activated T cells, lifetime and current suicidal behaviors, number of depressive episodes, and severity of depression, and 48.8% of its variance was explained by ACEs.

**Conclusions:** ACE-induced activation of T effector and cytotoxic cells and B cells with autoimmune potential, coupled with lowered Treg numbers are a key component of depression. The findings indicate that increasing ROI, the phenome of depression and suicidal behaviors, are caused by autoimmune processes, which are the consequence of ACEs and increasing sensitization of immune responses.

## Introduction

It has been established that major depressive disorder (MDD) is a neuro-immune disorder that exhibits various dysfunctions in the immune system. These dysfunctions include an elevation in immune-associated neurotoxicity, which leads to an increase in neuronal and astroglial toxicity. [1–5]. The latter, in turn, results in functional impairment of neuroaffective circuits [4]. The cytokine, macrophage-T lymphocyte, and immune response system (IRS) hypotheses of major depressive disorder (MDD) were formulated based on three lines of evidence from early research [2,3].

First, MDD has been found to be associated with the activation of T helper (Th)1 and macrophage M1 phenotypes. This is evidenced by elevated levels of pro-inflammatory cytokines or their soluble receptors. The cytokines and receptors that are being referred to are interleukin (IL)-1, IL-1 receptor antagonist (sIL-1RA), interferon (IFN)-γ, IL-2, IL-2 receptor (sIL-2R), IL-6, and tumour necrosis factor (TNF)-α [1,2,3]. Second, early research suggested that individuals with MDD exhibit a mild chronic inflammatory response, as evidenced by elevated levels of complement factors (such as C3), positive acute phase proteins (such as haptoglobin), and decreased levels of negative acute phase reactants (such as albumin and zinc) [5]. Third, the utilization of flow cytometry demonstrated the presence of T cell activation markers on peripheral blood mononuclear cells (PBMC) among individuals diagnosed with MDD. This was evidenced by a rise in the quantities or expression levels of CD4+, CD45+, CD25+, Ig+, and HLA-DR+ [6–8]. Those associations between MDD and the activation of the immune-inflammatory response system (IRS), Th1 and M1 activation, the acute phase or inflammatory response, and T cell activation have been established by recent reviews, systematic reviews, and meta-analyses [9–13].

In recent years, there has been an increase in knowledge and precision concerning the IRS theory of MDD. This has been achieved through the illustration of the participation of the compensatory immunoregulatory system (CIRS) in MDD [1,3,14]. There exists supporting evidence for the relative upregulation of anti-inflammatory or negative immuno-regulatory immune phenotypes, namely Th2 and Tregulatory (Treg), in individuals with MDD. This is demonstrated by heightened levels of IL-4, IL-10, and transforming growth factor (TGF)-β [3,15]. However, it has been observed that the acute phase of MDD is marked by a significant rise in the ratio of IRS/CIRS, leading to immune activation [3]. Furthermore, patients diagnosed with MDD demonstrate a reduction in Treg levels in comparison to healthy controls [16]. Furthermore, depressed individuals who have received treatment with antidepressants exhibit an increased abundance of CD4+CD25+FOXP3+Treg cells as compared to those who have not undergone any treatment for depression [17]. Studies utilizing animal models of depression have demonstrated an association between depression-like behaviors and diminished Treg functions [18]. The phenotypes of Th2 and Treg cells effectively curtail the deleterious activities of effector cells, thus facilitating immune equilibrium and homeostasis [3].

Recent studies conducted by Rachayon et al. [19] and Maes et al. [20] have provided a more precise identification of the cell surface markers that are associated with the increased IRS/CIRS ratio in individuals with acute MDD. The latter individuals demonstrate various immunological modifications, such as increased Th1 polarization and increased concentrations of IL-16 [21]. Furthermore, there was a rise in both quantity and expression of T cell activation indicators, specifically CD71+ and CD40L+, on T lymphocytes, predominantly CD3+ and CD4+ but also CD8+. On the other hand, a reduction in the manifestation of Treg cells was observed in depression, specifically those that include CD25+FoxP+CD152+ (CTLA-4 Tregs that have an inhibitory effect on the IRS), CD25+FoxP3+GERP+ (TGF-β secreting Tregs), and CD25+FoxP+CB1+ (cannabinoid receptor CB1) [19,20]. Moreover, there was a rise in the quantity and expression of B cells bearing the cannabinoid receptor CB2 (CD20+CB2+ cells) [20]. Increased IL-16 production and the activation of T cells have been proposed as possible explanations for this phenomenon, which may lead to detrimental autoimmune responses linked to MDD, particularly those mediated by IgM [20].

To summarize, the findings indicate that, in the acute stage of severe MDD, there is an activation of M1, Th1 polarization, activation of CD4 via IL-16, and consequent activation of CD8+ effector cells [3,19,20,21]. These events take place amidst reduced Treg functions, suggesting a disruption of peripheral immune tolerance. The heightened potential for neuroimmunotoxicity leading to damage to astroglial and neuronal projections, resulting in MDD, can be attributed to the augmented immunotoxicity caused by the production of M1, Th1, Th17, Th2 products, complement, and certain acute phase proteins, along with reduced Treg-associated neuroprotection [3,4,22].

However, it remains unclear whether heightened T cell activation and increased levels of CD20+CB2+ cells, as well as reduced Treg protection, are linked to adverse childhood experiences (ACE), the reoccurrence of depressice episodes, suicidal ideation, and suicidal attempts, referred to as recurrence of illness (ROI), and the current phnenome of depression, conceptualized as the first factor derived from the severity of depression, anxiety, and current suicidal behaviors. The authors employed the precision nomothetic approach [22–24], to create novel neuro-immune precision models. These models revealed that the effects of ACE on the depression phenotype were mediated through ROI and various ROI-associated pathways. These pathways included the production of neurotoxic cytokine profiles, decreased antioxidant defenses, and increased oxidative damage [22,23,25,26]. Importantly, two novel subgroups of MDD could be identified based on the severity of the phenotype, ROI, and those neuro-immune pathways. These subgroups are referred to as Major Dysmood Disorder (MDMD) and Simple Depression (SDMD). MDMD is distinguished by heightened depression severity, increased ROI, and a greater number of episodes and suicidal behavior [22,24,25,27].

The objective of the current study was to clarify the influence of ACE and ROI on the stimulation of T cells, Treg, and CB2+ expressing CD3+, CD4+, CD8+, and CD20+ B cells. Furthermore, the research aimed to investigate whether these lymphocyte populations mediate the effects of ACE on the phenome of depression, and whether the aberrations in the lymphocyte populations are more prominent in MDMD compared to SDMD.

### Methodology Participants

We conducted this research with MDD outpatients at Chulalongkorn Hospital’s psychiatric department in Bangkok, Thailand. Patients between the ages of 18 and 65 who spoke Thai, received a DSM-5 diagnosis of MDD from an experienced psychiatrist, and scored at least 17 on the Hamilton Depression Rating Scale (HAM-D) [28] were considered for inclusion. Posters and word of mouth were used to find healthy volunteers in the same area (Bangkok, Thailand) as the patients. Patients who met the criteria for bipolar, substance use, schizo-affective, post-traumatic stress, psycho-organic, and obsessive-compulsive disorder, or schizophrenia, were not included. Controls were omitted if they met the criteria for any axis 1 DSM-5 [29] condition or had a family history of MDD or bipolar disorder (BD). Healthy controls and patients were not included if they had any of the following conditions: neurological, neurodegenerative, or neuroinflammatory diseases like multiple sclerosis, epilepsy, Alzheimer’s, or Parkinson’s; (auto)immune diseases like COPD, inflammatory bowel disease, cancer, asthma, type 1 diabetes, Long COVID, COVID-19 infection (lifelong), or rheumatoid arthritis; or a history of allergic or inflammatory responses within the previous three months. Participants who had recently taken nonsteroidal anti-inflammatory drugs (NSAIDs) were also disqualified, as were those who had taken therapeutic levels of antioxidants or omega-3 polyunsaturated fatty acid supplements in the three months before the trial. Pregnant or nursing women were not permitted to take part. Benzodiazepines were used by 22 patients, sertraline by 18 patients, other antidepressants (including fluoxetine, venlafaxine, escitalopram, bupropion, and mirtazapine) by 8, atypical antipsychotics by 14, and mood stabilizers by 4. The potential effects of these pharmacological variables were included in the statistical analysis.

All subjects and controls gave their informed written consent to take part in the study before their participation. Both international and Thai privacy and ethics rules were followed during the study’s execution. International guidelines for the protection of human research subjects, such as the Declaration of Helsinki, the Belmont Report, and the International Conference on Harmonization in Good Clinical Practice, were followed in approving the study (#528/63) by the Chulalongkorn University Faculty of Medicine in Bangkok, Thailand.

### Clinical measurements

An expert psychiatrist administered the 17-item Hamilton Depression Rating Scale (HAMD) to assess the severity of depressive symptoms [28]. The same psychiatrist employed the Mini-International Neuropsychiatric Interview (M.I.N.I.) to make a diagnosis of axis-1 mental disease [30]. The Columbia-Suicide Severity Rating Scale (C-SSRS) lifeline version [31] was used to assess the extent to which the participants suffered from suicidal ideation or had committed suicide attempts. The diagnosis of MDD was made using the DSM-5 criteria [29]. In addition, we categorized the patients into MDMD (the most severe subtype) and SDMS (milder depression) according to previously published criteria [22]. Self-reported levels of state anxiety were measured using the Thai translation of the State-Trait Anxiety Inventory, state version [32]. Demographic information, such as age, gender, medical and psychiatric history, and number of depressive episodes, was gathered through semi-structured interviews done by a professional research assistant.

The Adverse Childhood Experiences (ACE) Questionnaire, consisting of 28 items that score 10 domains, was utilized to evaluate ACEs [33]. These domains include mental trauma, physical trauma, sexual abuse, mental neglect, physical neglect, witnessing domestic violence, having a family member with drug abuse, having a family member with depression/mental illness, experiencing parental separation, death, or divorce, and having a family member who is incarcerated. We computed the total ACE score (labelled: ACEsum) using the scores (yes/no) on the 10 domains. In addition, we computed a principal component (PC) score using the raw scores on mental trauma, physical trauma, mental neglect, domestic violence, a family history of mental disease, or loss of a parent, labeled PC_ACE [25]. As explained by Maes et al. [25], lifetime suicidal behaviors (LT_SB) were computed as the first PC extracted from 11 lifetime C-SSRS items on suicidal ideation (labeled: LT_SI) and lifetime attempts (Labeled LT_SA). Consequently, a composite was computed using both SI and SA, labeled LT_SB. Consequently, ROI was calculated as the first PC extracted from the number of depressive episodes, the C-SSRS item “lifetime suicidal ideation”, the C-SSRS item on number of actual lifetime suicidal attempts, and LT_SB. Current suicidal behaviors (“current_SB”) were computed employing 9 C-SSRS items that assessed current (last month) complaints of suicidal ideation and attempts. Using the total HAMD score, the total STAI score, and the current_SB score, the first PC was determined to characterize the “phenome of depression”, labeled PC_phenome [25]. Weight (in kilos) was divided by height (in square meters) to determine body mass index (BMI). Tobacco use disorder was identified according to DSM-5 criteria.

### Assays

Between 8 and 10 a.m. in the morning, 20 mL of fasting blood was collected using BD Vacutainer® EDTA (10 mL) and BD Vacutainer® SSTTM (5 mL) vials (BD Biosciences, Franklin Lakes, NJ, USA). Blood was allowed to coagulate at ambient temperature for 30 minutes to extract serum. The containers were centrifuged for 10 minutes at 4 degrees Celsius and 1100 g to obtain serum, which was subsequently frozen at −80 degrees Celsius. PBMCs were isolated using Ficoll® Paque Plus (GE Healthcare Life Sciences, Pittsburgh, PA, USA) in a density gradient centrifugation procedure (30 minutes at 900 g). Hemocytometers were used to enumerate the number of cells, and trypan blue, 0.4% solution, pH 7.2-7.3 (Sigma-Aldrich Corporation, St. Louis, MO, USA) was used to determine the percentage of viable cells. Upon counting both total and blue stained cells under various conditions, it was determined that more than 95% of cells were alive.

Anti-human CD3 antibody (OKT3, from eBioscience) was coated at 5 µg/mL on 96-well plates for the evaluation of PBMC stimulation the following day. Each well was injected with 3 x 10^5^ PBMCs and 5 µg/mL anti-human CD28 antibody (CD28.2, eBioscience). To culture the cells, 1% penicillin-streptomycin solution (Gibco Life Technologies, Carlsbad, CA, USA) and 10% fetal bovine serum (FBS) were added to RPMI 1640 containing L-glutamine. The cells were allowed to proliferate at 37 degrees Celsius and 5% carbon dioxide for three days. As a negative control, PBMCs were grown without anti-CD3 and anti-CD28 antibodies for the same amount of time. Consequently, after 72 hours of incubation, samples can be categorized as either unstimulated or stimulated with anti-CD3/CD28 beads. We labeled the 3×10^5^ PBMCs with monoclonal antibodies for 30 minutes to delineate different lymphocyte phenotypes, including CD3-PEcy7, CD4-APCcy7, CD8-APC, CD20-FITC, CB2-AF700, CD40L-FITC, CD69-AF700, CD71-PerCPcy5.5, HLA-DR-PE594, CD25-APC, CD152 PE Dazzle594, GARP-PE, CB1-AF700 and 7-AAD (Biolegend, BD Biosciences and R&D Systems). For Treg cells, we first stained cells with surface markers (CD3, CD4, CD25), then performed intracellular FoxP3 staining using the FoxP3/Transcription Factor Staining Buffer Set (eBioscience) for fixation and permeabilization prior to staining with anti-Foxp3-FITC antibody (Biolegend). Immediately following staining, at least 50,000 lymphocytes were counted using size and granularity criteria by flow cytometry. Our gating techniques for evaluating the lymphocyte phenotypes were displayed in our previous papers [19,20]. We employed the LSRII flow cytometer (BD Biosciences) for the flow cytometry, and FlowJo X (Tree Star Inc., Ashland, OR, USA) to perform data analysis. Consequently, we counted the following cells: CD3+CB2+, CD4+CB2+, CD8+CB2+, CD20+CB2+, CD25+FoxP3+CB1+, CD25+FoxP3+CD152+, CD25+FoxP3+GARP+, CD3+CD71+, CD3+CD40L+, CD3+HLADR+, CD4+CD71+, CD4+CD40L+, CD4+HLADR+, CD8+CD71+, CD8+CD40L+, and CD8+HLADR+.

### Statistics

We employed Pearson’s product moment correlation or Spearman’s rank order correlation coefficients for the purpose of examining the relationships between scale variables. Among categories, we compared nominal variables via analysis of contingency tables (χ2-test) and continuous variables via analysis of variance (ANOVA). We utilized transformations such as logarithmic, square root, rank-based inversed normal (RINT), and Winsorization when necessary to normalize the data distribution of the indicators. Using manual multiple regression analysis, the effects of explanatory variables (e.g., ACE and ROI) on dependent variables (immune profiles) were evaluated. In addition, we utilized an automatic forward stepwise regression strategy with a p-to-enter of 0.05 and a p-to-remove of 0.06 to determine which variables would be included and which would be excluded from the final regression model. Residuals, residual plots, and data quality were always evaluated in the final model. Using the modified Breusch-Pagan test and the White test, heteroskedasticity was investigated. We examined the probability of multicollinearity and collinearity utilizing the tolerance (cut-off value: 0.25) and variance inflation factor (cut-off value: > 4), as well as the condition index and variance proportions from the collinearity diagnostics table. We estimated the standardized β coefficients using t-statistics and exact p-values, F-statistics (and p-values), and the total variance as the effect size (partial Eta squared). We also computed partial regression analyses, including partial regression graphs, based on the results of the linear regression analyses. A first pre-specified repeated measures generalized estimating equation (GEE) was used to assess the differences between controls, SDMD, and MDMD, as well as the effects of stimulation (unstimulated versus stimulation), and the interaction between diagnosis by stimulation with anti-CD3/CD28 beads, while allowing for the effects of sex, age, TUD, BMI, and drug state of the patients. Another pre-specified (repeated measures) GEE was performed with fixed categorical effects of the condition (unstimulated versus stimulated) and ACE (total sum or subdomain scores), while accounting for the effects of the confounding variables. There were no missing variables in any of the cell populations or clinical data. The statistical significance threshold for two-tailed tests was established at p<0.05. To conduct the statistical analyses, we utilized IBM’s Windows version of SPSS 28.

Multiple regression analysis, which computes how immune cell populations might predict the depressive phenome, is the primary statistical analysis. A priori power analysis (G*Power 3.1.9.4) for a linear multiple regression recommends that the minimum sample size should be 48 given an effect size of 0.25 (equal to 20% explained variance), power=0.8, alpha=0.05, and 3 covariates.

The precision nomothetic models were constructed using Partial Least Squares (PLS) Analysis, which has been expounded upon in prior literature [34–38]. The present study employed SmartPLS path analysis [34,35] to evaluate the causal associations among ACE, ROI, T/B cell phenotypes, and the depression phenome. The study employed a complete PLS analysis, which was executed only after ensuring that the outer and inner models satisfied predetermined quality criteria. These criteria included: a) all latent vectors exhibiting an average variance extracted (AVE) greater than 0.50, Cronbach’s alpha, and composite reliabilities (rho a and rho c) scores greater than 0.7 and 0.8, respectively; b) all loadings on the latent vectors exceeding 0.65 at p<0.001; c) the models being reflective models, as specified by Confirmatory Tetrad Analysis; d) the model fit SRMR being less than 0.08; e) the establishment of discriminant validity through the Heterotrait-Monotrait ratio (HTMT); and e) adequate model replicability, as confirmed by PLSpredict and Q2 predict, and the cross-validated predictive ability test (CVPAT) [34–38]. A complete PLS analysis was conducted using 5,000 bootstrap samples to ascertain the total direct and indirect effects, specific indirect effects, and path coefficients, along with their precise p-values.

## Results

**Table 1** shows the demographic and clinical data of the participants in controls and patients with MDMD and SDMD. There were no significant differences in age, sex, education, or TUD among the study groups. The BMI was somewhat higher in patients than controls. Nevertheless, we have controlled all data for the effects of age, sex, BMI, TUD, education, and drug state. The HAMD, STAI, and ACE scores were higher in patients than in controls, while LT_SB, number of episodes, Current_SB, and PC_phenome were significantly higher in MDMD than in SDMD.

**Table 1.**
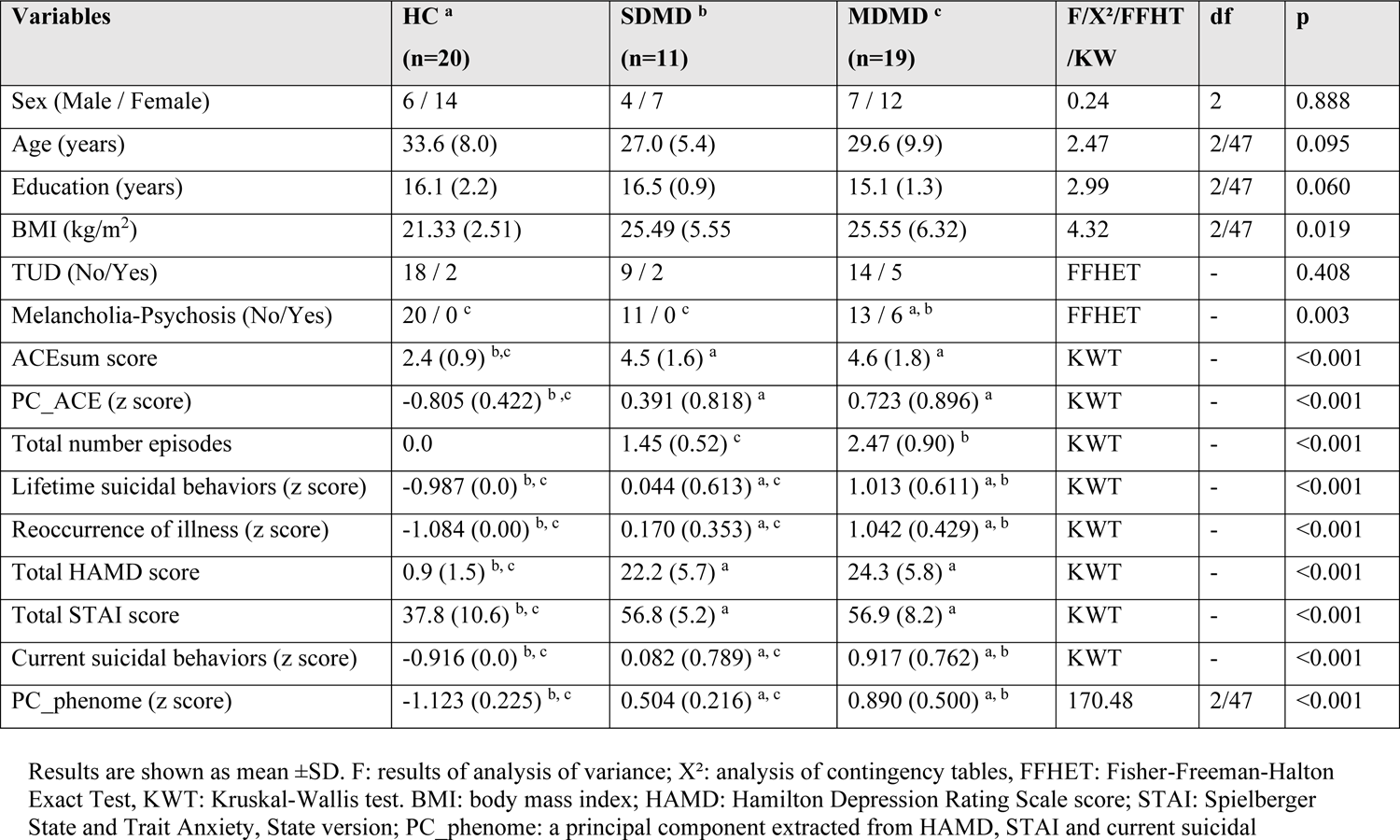
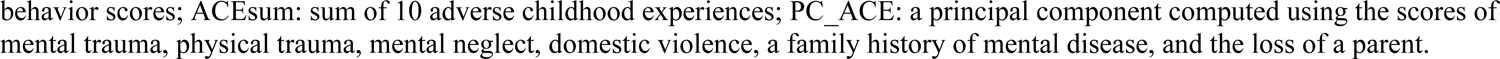
Demographic and clinical data of the healthy controls (HC) and depressed patients divided into those with simple depression (SDMD) and Major Dysmood Disorder (MDMD)

| Variables | HC <sup>a</sup><br>(n=20) | SDMD <sup>b</sup><br>(n=11) | MDMD <sup>c</sup><br>(n=19) | F/X<sup>2</sup>/FFHT<br>/KW | df | p |
| --- | --- | --- | --- | --- | --- | --- |
| Sex (Male / Female) | 6 / 14 | 4 / 7 | 7 / 12 | 0.24 | 2 | 0.888 |
| Age (years) | 33.6 (8.0) | 27.0 (5.4) | 29.6 (9.9) | 2.47 | 2/47 | 0.095 |
| Education (years) | 16.1 (2.2) | 16.5 (0.9) | 15.1 (1.3) | 2.99 | 2/47 | 0.060 |
| BMI (kg/m<sup>2</sup>) | 21.33 (2.51) | 25.49 (5.55) | 25.55 (6.32) | 4.32 | 2/47 | 0.019 |
| TUD (No/Yes) | 18 / 2 | 9 / 2 | 14 / 5 | FFHET | - | 0.408 |
| Melancholia-Psychosis (No/Yes) | 20 / 0 <sup>c</sup> | 11 / 0 <sup>c</sup> | 13 / 6 <sup>a, b</sup> | FFHET | - | 0.003 |
| ACEsum score | 2.4 (0.9) <sup>b, c</sup> | 4.5 (1.6) <sup>a</sup> | 4.6 (1.8) <sup>a</sup> | KWT | - | <0.001 |
| PC_ACE (z score) | -0.805 (0.422) <sup>b, c</sup> | 0.391 (0.818) <sup>a</sup> | 0.723 (0.896) <sup>a</sup> | KWT | - | <0.001 |
| Total number episodes | 0.0 | 1.45 (0.52) <sup>c</sup> | 2.47 (0.90) <sup>b</sup> | KWT | - | <0.001 |
| Lifetime suicidal behaviors (z score) | -0.987 (0.0) <sup>b, c</sup> | 0.044 (0.613) <sup>a, c</sup> | 1.013 (0.611) <sup>a, b</sup> | KWT | - | <0.001 |
| Reoccurrence of illness (z score) | -1.084 (0.00) <sup>b, c</sup> | 0.170 (0.353) <sup>a, c</sup> | 1.042 (0.429) <sup>a, b</sup> | KWT | - | <0.001 |
| Total HAMD score | 0.9 (1.5) <sup>b, c</sup> | 22.2 (5.7) <sup>a</sup> | 24.3 (5.8) <sup>a</sup> | KWT | - | <0.001 |
| Total STAI score | 37.8 (10.6) <sup>b, c</sup> | 56.8 (5.2) <sup>a</sup> | 56.9 (8.2) <sup>a</sup> | KWT | - | <0.001 |
| Current suicidal behaviors (z score) | -0.916 (0.0) <sup>b, c</sup> | 0.082 (0.789) <sup>a, c</sup> | 0.917 (0.762) <sup>a, b</sup> | KWT | - | <0.001 |
| PC_phenome (z score) | -1.123 (0.225) <sup>b, c</sup> | 0.504 (0.216) <sup>a, c</sup> | 0.890 (0.500) <sup>a, b</sup> | 170.48 | 2/47 | <0.001 |

Results are shown as mean ± SD. F: results of analysis of variance; X<sup>2</sup>: analysis of contingency tables, FFHET: Fisher-Freeman-Halton Exact Test, KWT: Kruskal-Wallis test. BMI: body mass index; HAMD: Hamilton Depression Rating Scale score; STAI: Spielberger State and Trait Anxiety, State version; PC\_phenome: a principal component extracted from HAMD, STAI and current suicidal
behavior scores; ACEsum: sum of 10 adverse childhood experiences; PC\_ACE: a principal component computed using the scores of mental trauma, physical trauma, mental neglect, domestic violence, a family history of mental disease, and the loss of a parent.

### Lymphocytes profiles in MDMD and SDMD

**Table 2** shows the results of GEE, repeated measures, which analyzed the associations between the lymphocyte phenotypes and the diagnosis (MDMD and SDMD and controls). This table shows the results of interaction patterns between diagnosis by condition (unstimulated versus stimulated) and the measurement of the immune cells in both conditions when the interaction term is significant. If not, we show the statistical results of the comparisons among the three subgroups. The stimulated numbers of the immune cells were always higher than the unstimulated levels (all p<0.001). We found that CD20+CB2+ was significantly higher in MDMD than in controls and SDMD. The percentages of baseline CD3+CB2+ cells and CD25+FoxP3+CB1+ were significantly lower in SDMD than in the other 2 groups, while baseline CD4+CB2+ % was lower in SDMD than in MDMD. The unstimulated CD25+FoxP3+GARP+ % was significantly lower in SDMD than in controls. The baseline percentages of all FoxP3+ cells combined were significantly lower in MDD as compared with controls, while the stimulated levels were higher in MDMD than in the two other groups. The baseline levels of CD3+CD71+ % and CD4+CD71+ % were higher in MDMD than in the two other groups. Baseline levels of CD3+CD40L+, CD4+CD40L+, CD4+HLADR+, and CD8+CD40L+ % were higher in MDMD than in controls.

**Table 2.**
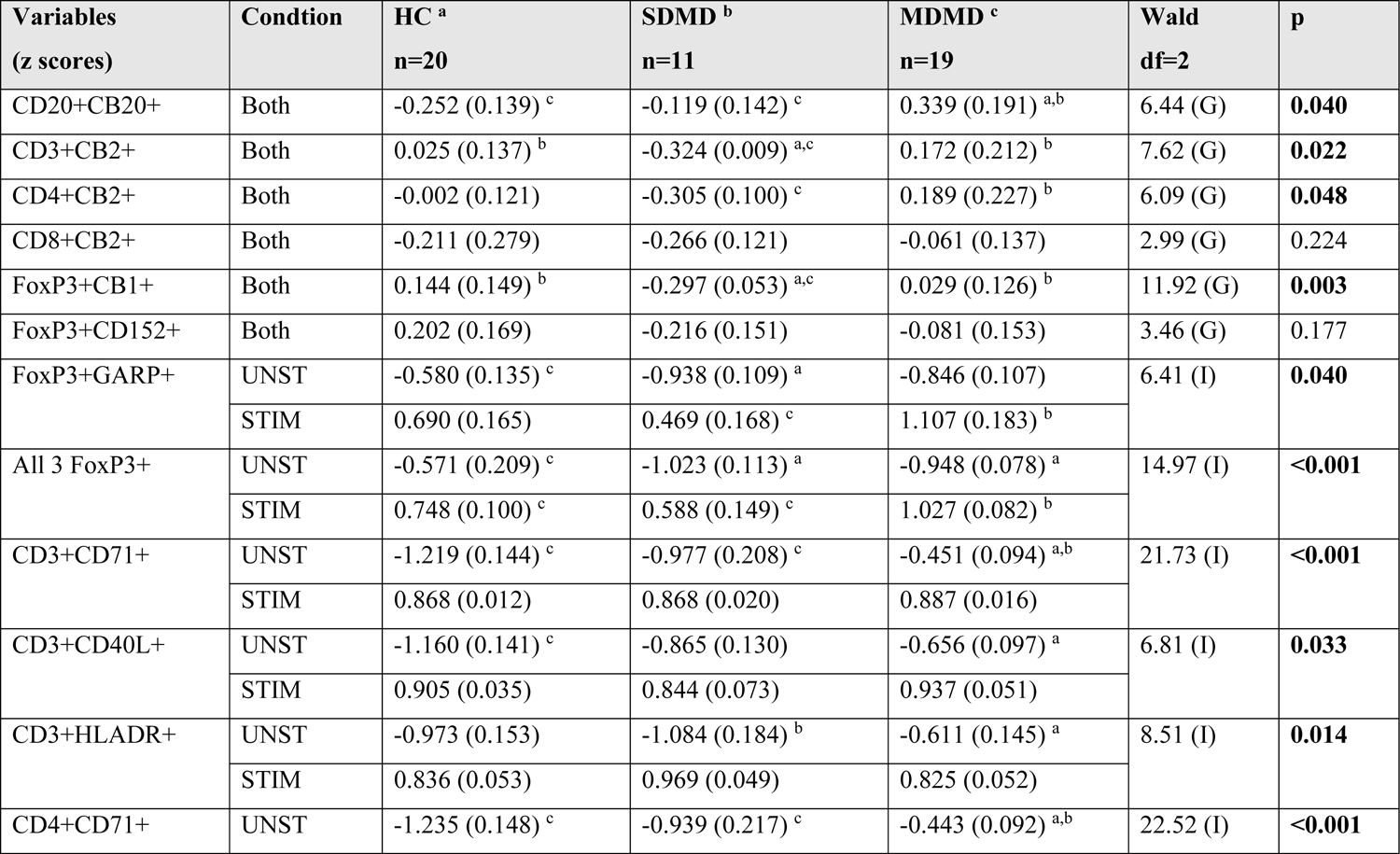
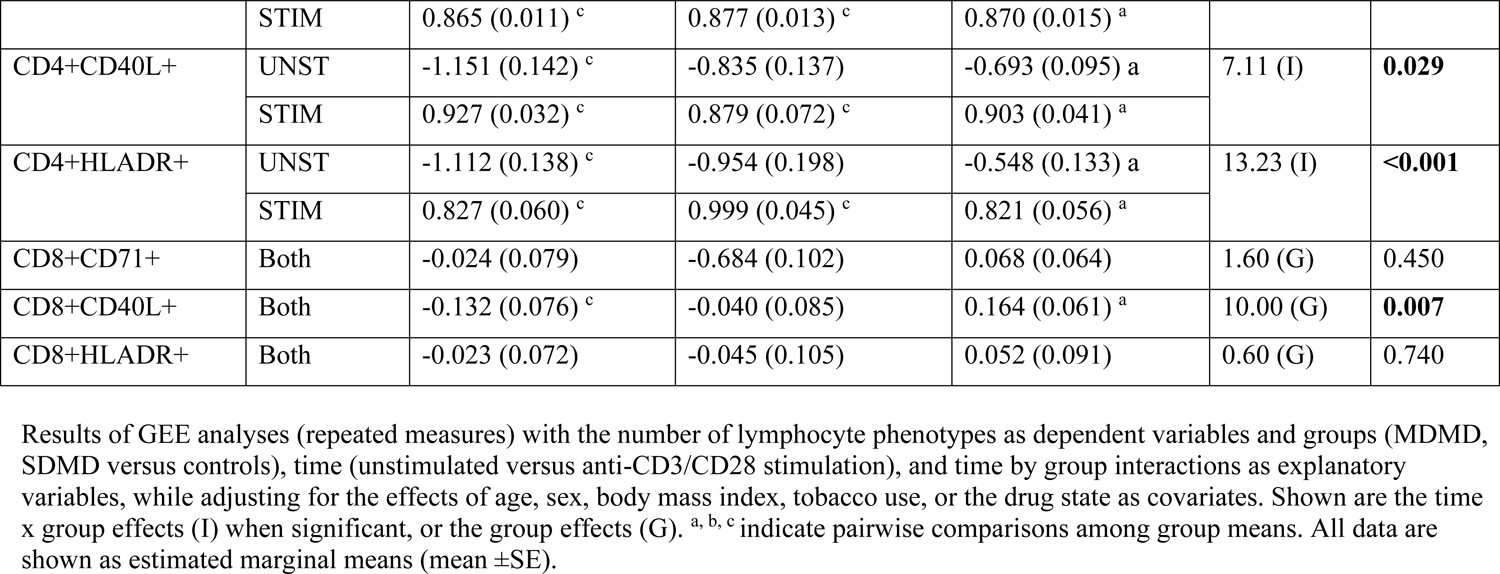
Differences in unstimulated (UNST) and anti CD3/CD28 bead stimulated (STIM) numbers of lymphocyte phenotypes in healthy controls (HC) and depressed patients, divided into those with simple depression (SDMD) and Major Dysmood Disorder (MDMD)

### Lymphocyte phenotypes and depression phenome severity

To decipher the effects of the lymphocyte phenotypes on the phenome we have performed PLS analyses. **Figure 1** shows a first PLS analysis that examined the combined effects of the lymphocytes on the phenome. The latter was conceptualized as a latent vector (LV) extracted from the HAMD, STAI, and current_SB. Direct predictors were the stimulated CD20+CB2+ % (entered as a single indicator), Treg (entered as a LV extracted from CD25+FoxP3+CB1+, CD25+FoxP3+CD152+, and CD25+FoxP3+GARP+) and T cell activation (entered as a LV extracted from CD3CD71+, CD3+CD40L+, CD4+CD71+, CD4+CD40L+, and CD4+HLADR+cells). All three constructs (Phenome, Treg, T cell activation) showed adequate psychometric qualities, namely AVE (0.783, 0.660 and 0.794, respectively), Cronbach’s alpha (0.861, 0.749, and 0.935, respectively), rho a (0.865, 0.829 and 0.947, respectively) and rho c (0.915, 0.851 and 0.947, respectively). The overall model fit was sufficient (SRMR=0.073), and the model shows adequate replicability (Q2 predict values are all > 0.180, PLS versus indicator average: t=2.79, p=0.008). This first PLS model shows that 40.8% of the variance in the phenome was explained by CD20+CB2+, T cell activation (both positively) and Treg (negatively).

**Figure 1.**
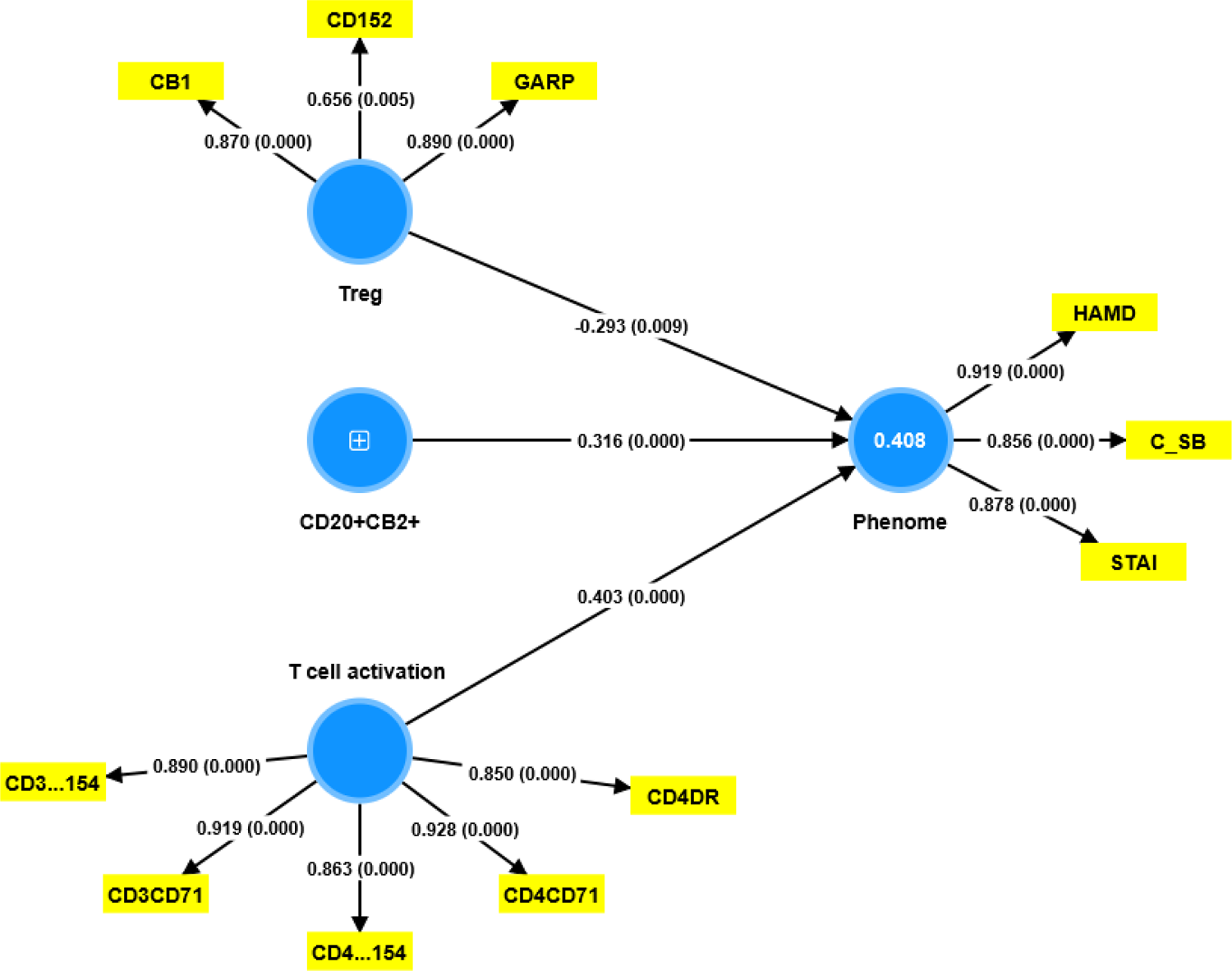
Results of Partial Least Squares (PLS) Analysis. This model examines the combined effects of CD20+CB2+ % (entered as a single indicator), baseline Treg (entered as a latent vector (LV) extracted from CD25+FoxP3+CB1+, CD25+FoxP3+CD152+, and CD25+FoxP3+GARP+), and T cell activation (entered as a LV extracted from CD3+CD71+, CD3+CD40L+, CD4+CD71+, CD4+CD40L+, and CD4+HLADR+ cells). The phenome of depression was conceptualized as a factor extracted from the Hamilton Depression Rating Scale (HAMD), Spielberger State and Trait Anxiety, State Version (STAI), and current suicidal behavior (C_SB) scores. The results of complete PLS analysis are shown as pathway coefficients (p-values) in the inner model, and loadings (p values) in the outer model; the figure in the blue circle indicates explained variance.

**Table 3** shows the T cell activation, CD20+CB2+, and Treg latent variable scores (see Figure 1) as well as the z unit-based composites (deduced from Figure 1) z Tcell activation + z CD20+CB2 (zT + zBCB2) and zT + zBCB2 – zTreg in the three study groups. T cell activation and CD20+CB2+ were significantly higher in MDMD than in controls, and zT + zBCB2 higher in MDMD than in controls and SDMD. Treg was significantly lower in SDMD than in controls, while zT + zBCB2 - zTreg was significantly increased in both MDD groups.

**Table 3.**
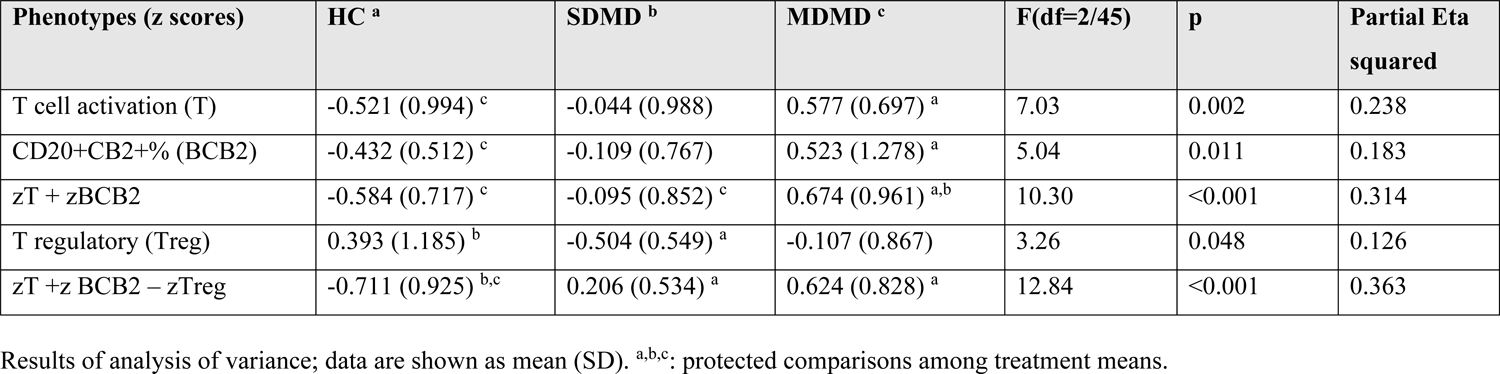
Difference in lymphocyte profiles between healthy controls (HC), major dysmood disorder (MDMD), and simple dysmood disorder (SDMD)

### Associations lymphocyte phenotypes and ACE and ROI

**Table 4** shows the correlation matrix between the baseline levels of the lymphocyte phenotypes (except CD20+CB2+, which is shown as the stimulated values). The ACEsum score was significantly associated with CD20+CB2+, CD3+CB2+, CD3+CD40L+, and CD4+40L+ cells. The number of episodes, LT_SB, ROI, and PC_phenome was significantly associated with CD20+CB2+, CD3+CD71+, CD3+CD40L+, CD4+CD71+, CD4+CD40L+, and CD4HLADR+cells. In addition, the number of episodes and ROI were also associated with the CD8+CD40L+ cells. There were significant associations between ACEsum and T cell activation, Treg, zT+zBCB2, and zT + zBCB2-Treg. LT_SB, ROI, and PC_phenome were significantly associated with the same indices, except with Treg. **Figures 2** and **Figure 3** show the partial regression of the zT + zBCB2 - zTreg on ACEsum and ROI scores, respectively (age-sex adjusted). **Figure 4** shows the partial regressions of the depression phenome on zT + zBCB2-zTreg (adjusted for age, sex). In the restricted study group of depressed patients, we found significant Spearman rank order correlations between ROI and T cell activation (r=0.508, p=0.002, n=29, one-tailed), CD20+CB2+ (r=0.334, p=0.038), zT + zBCB2 (r=0.576, p<0.001) and zT + zBCB2 - zTreg (r=0.525, p=0.002).

**Figure 2.**
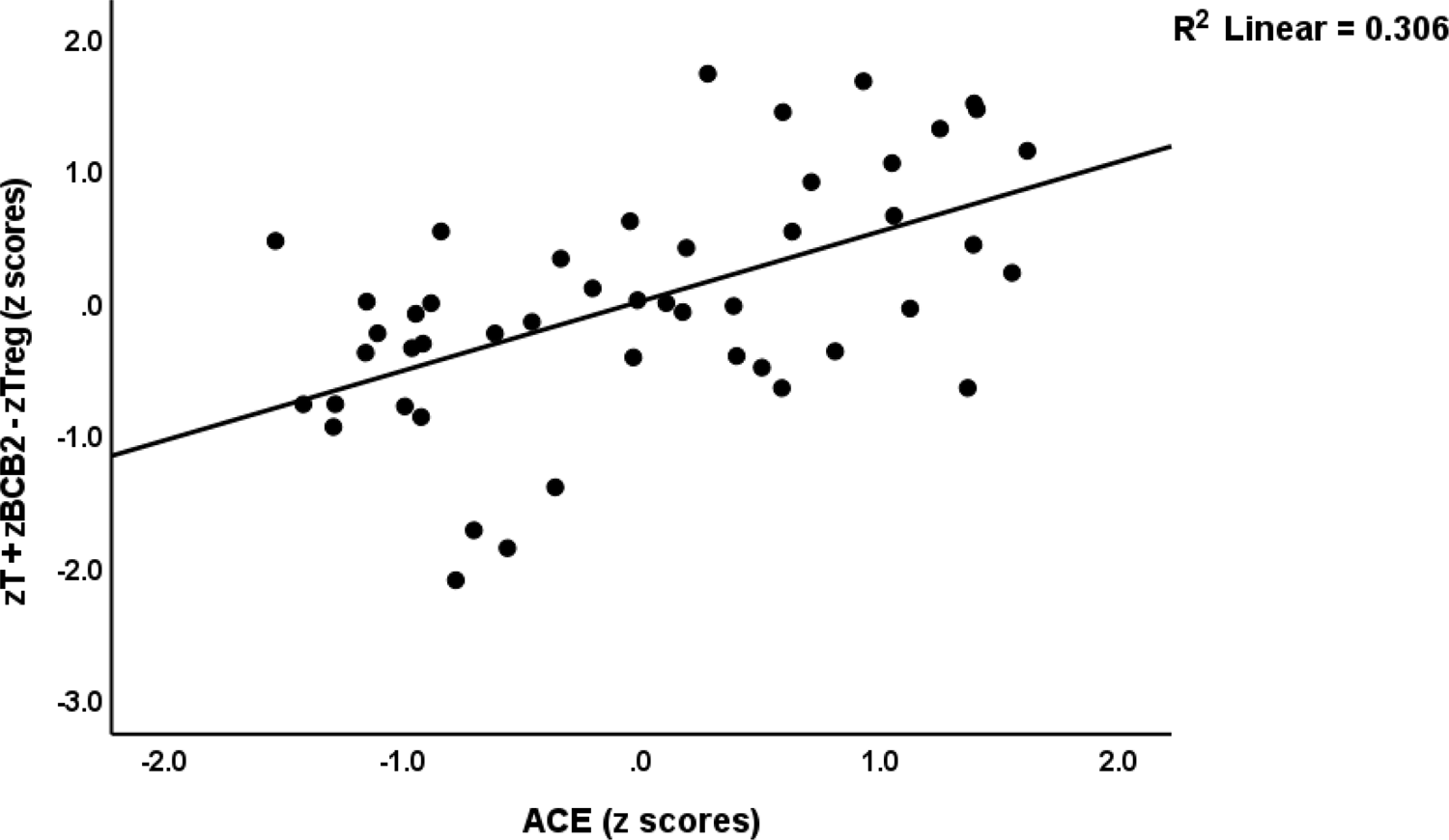
Partial regression of the ratio of T cell activation + increased CD20+CB2+ B cells / regulatory cells (zT + zBCB2 – zTreg) on the sum of adverse childhood experiences (t=4.30, p<0.001).

**Figure 3.**
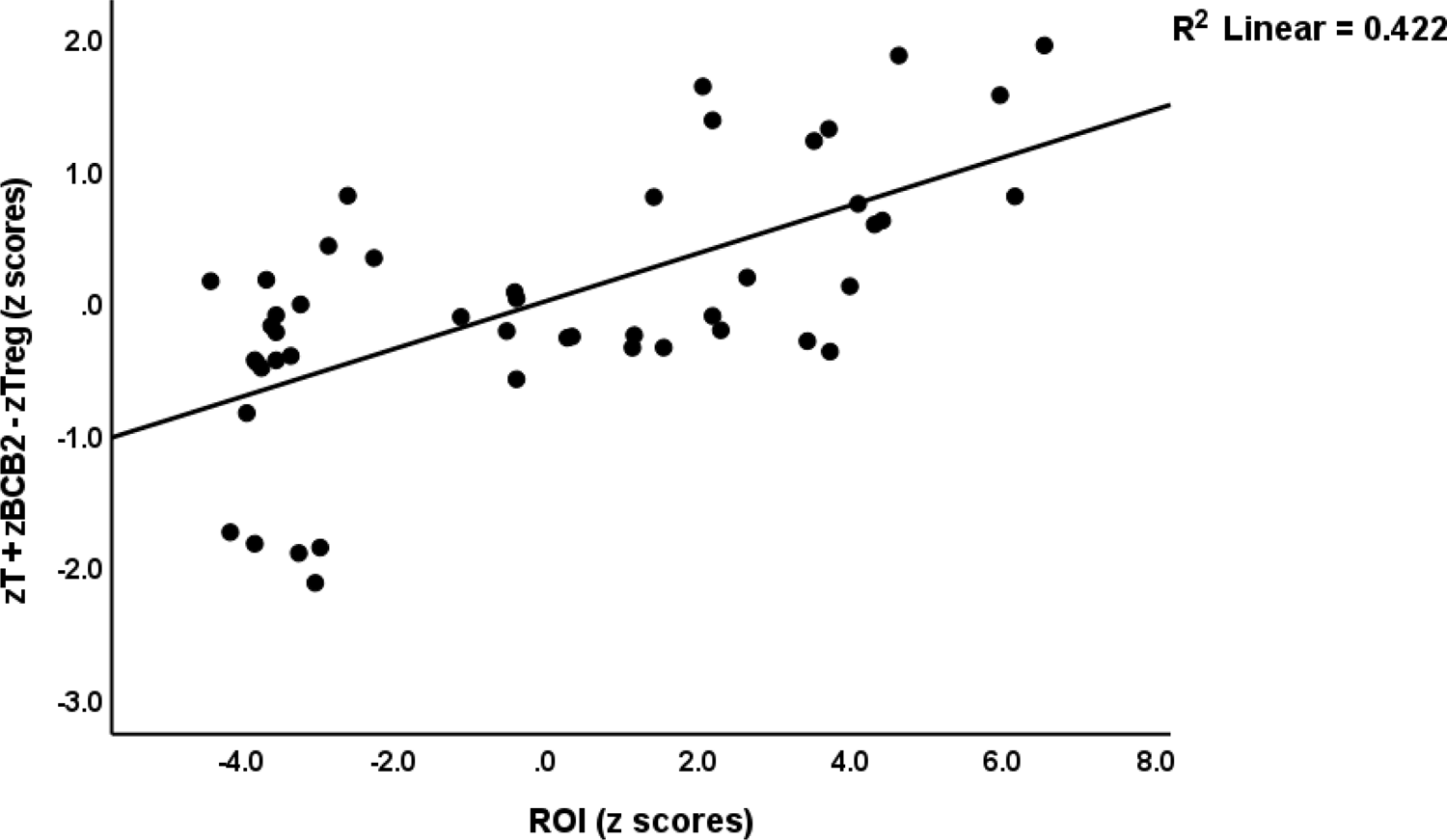
Partial regression of the ratio of T cell activation + increased CD20+CB2+ B cells / regulatory cells (zT + zBCB2 – zTreg) on the recurrence of illness (t=5.67, p<0.001).

**Figure 4.**
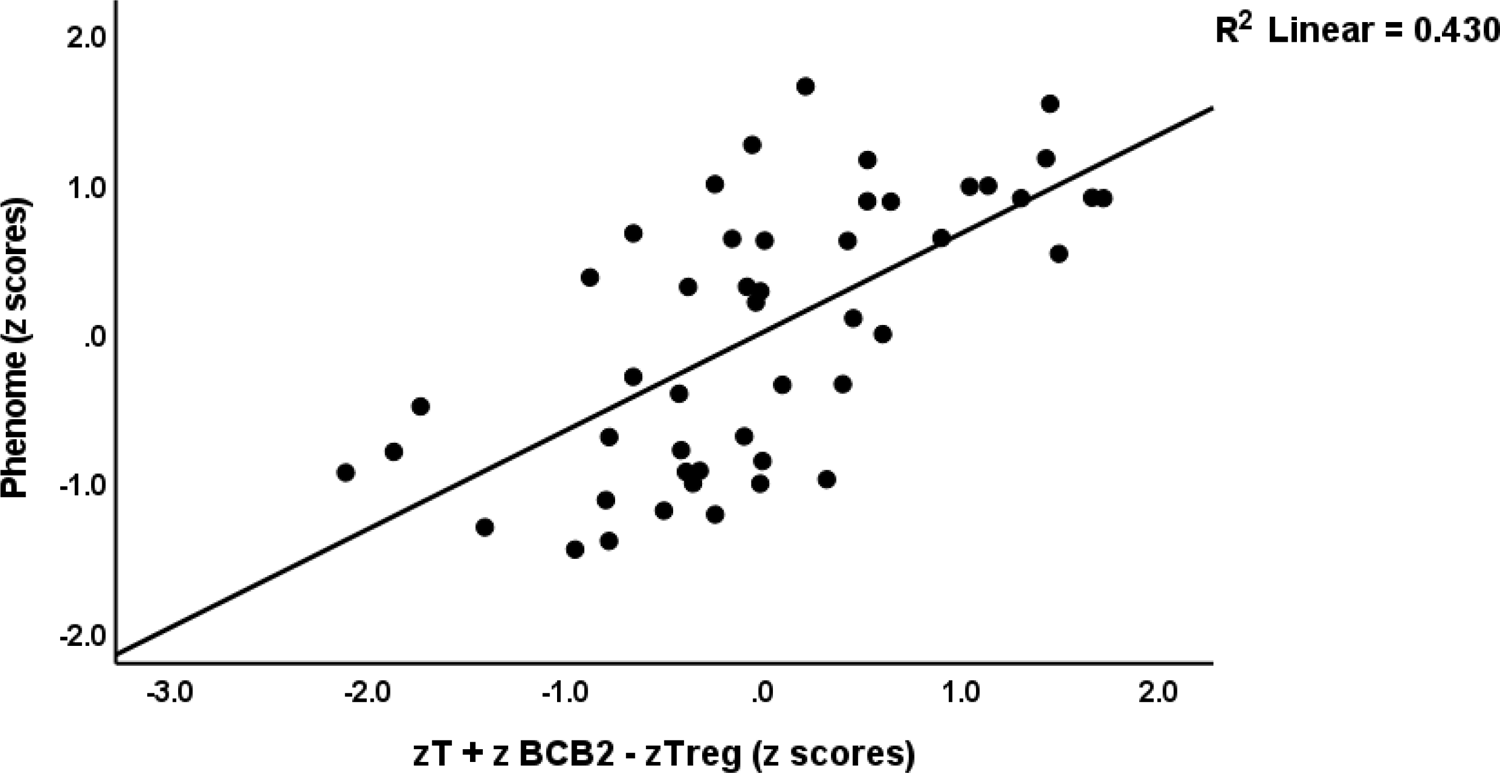
Partial regression of the depression phenome on the ratio of T cell activation + increased CD20+CB2+ B cells / regulatory cells (zT + zBCB2 – zTreg) (t=5.63, p<0.001).

**Table 4.** Intercorrelation matrix between the lymphocyte phenotypes and clinical features of depression

| Phenotypes (%) | ACEsum | #episodes | LT_SB | ROI | PC_phenome |
| --- | --- | --- | --- | --- | --- |
| CD20+CB2+* | 0.404 ** | 0.417 ** | 0.442 ** | 0.452 ** | 0.442 * |
| CD3+CB2+ | 0.298 * | 0.025 | -0.022 | 0.016 | -0.033 |
| CD4+CB2+ | 0.180 | -0.014 | -0.120 | -0.041 | 0.021 |
| CD8+CB2+ | 0.160 | -0.182 | -0.055 | -0.146 | -0.106 |
| FoxP3+CB1+ | -0.197 | -0.198 | -0.098 | -0.128 | -0.237 |
| FoxP3+CD152+ | -0.154 | -0.256 | -0.063 | -0.166 | -0.148 |
| FoxP3+GARP+ | -0.203 | -0.263 | -0.153 | -0.204 | -0.237 |
| CD3+CD71+ | 0.115 | 0.522 ** | 0.433 ** | 0.533 ** | 0.591 ** |
| CD3+CD40L+ | 0.302 * | 0.420 ** | 0.325 ** | 0.445 ** | 0.434 ** |
| CD3+HLADR+ | -0.009 | 0.268 | 0.227 | 0.265 | 0.266 |
| CD4+CD71+ | 0.137 | 0.525 ** | 0.436 ** | 0.537 ** | 0.626 ** |
| CD4+CD40L+ | 0.301 * | 0.374 ** | 0.310 * | 0.412 ** | 0.397 ** |
| CD4+HLADR+ | 0.076 | 0.375** | 0.375 ** | 0.405 ** | 0.431 ** |
| CD8+CD71+ | 0.0120. | 0.143 | 0.063 | 0.112 | 0.085 |
| CD8+CD40L+ | 0.117 | 0.390 ** | 0.186 | 0.341 * | 0.278 |
| CD8+HLADR+ | -0.037 | 0.189 | 0.126 | 0.154 | 0.096 |
| T cell activation (T) | 0.280 * | 0.523 ** | 0.500 ** | 0.590 ** | 0.566 ** |
| T regulatory (Treg) | -0.244 * | -0.241 * | -0.093 | -0.161 | -0.222 |
| zT + zBCB2 | 0.425 ** | 0.558 ** | 0.518 ** | 0.616 ** | 0.587 ** |
| zT +z BCB2 / zTreg | 0.478 ** | 0.661 ** | 0.526 ** | 0.681 ** | 0.643 ** |

\* P<0.05, \*\* P<0.01
All results of Spearman's rank order correlation coefficients

Furthermore, in depression, ACEs were significantly associated with lymphocyte profiles; for example, PC_ACE with CD20+CB2+ % (r=0.356, p=0.029), zT + zBCB2 (r=0.349, p=0.032) and zT +zBCB2 - zTreg (r=0.316, p=0.047).

**Figure 5** builds on the first PLS model (Figure 1) but adds the effects of ROI and ACEs, while the lymphocyte phenotypes are allowed to mediate their effects on the phenome. The same three validated LVs, as shown in Figure 1, were entered, as well as ROI (a LV extracted from number of episodes, LT_SA and LT_SI) and ACE (LV extracted from mental neglect, mental trauma, and physical trauma). Both LVs showed adequate psychometric properties, including AVE (0.740 and 0.647, respectively), Cronbach’s alpha (0.824 and 0.732), rho a (0.857 and 0.766, respectively), and rho c (0.895 and 0.846). With an SRMR of 0.075, the model fit was sufficient. The results show that 31.7% of the variance in T cell activation was explained by the ROI and ACE latent vectors, and that 21.6% of the variance in stimulated CD20+CB2+ was explained by ROI. In addition, there was a significant specific indirect effect of ACE on CD20+CB2+ % that was mediated by ROI (t=3.32, p=0.001).

**Figure 5.**
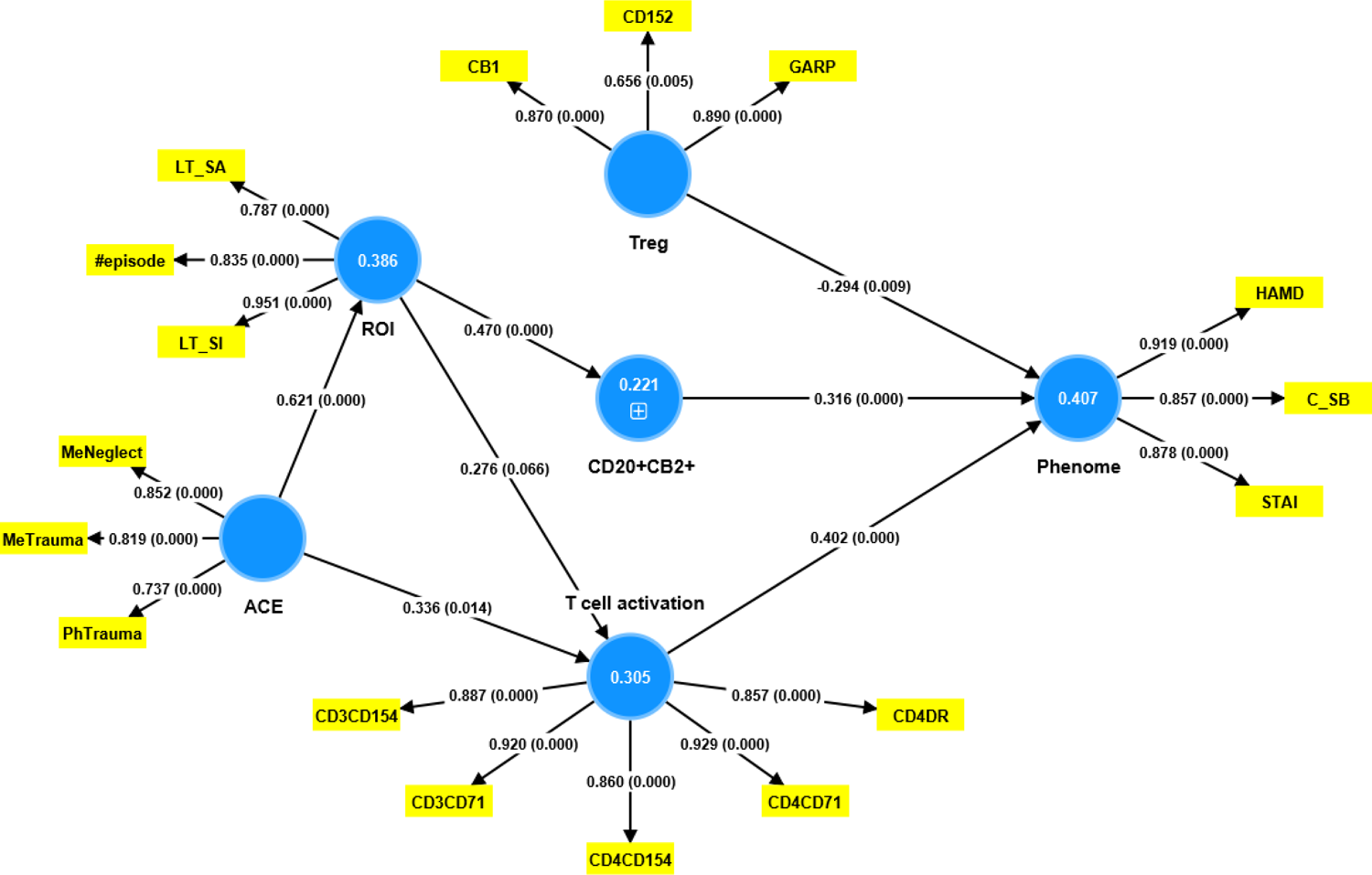
Results of Partial Least Squares (PLS) Analysis. This model examines the combined effects of CD20+CB2+ % (entered as a single indicator), baseline Treg, and T cell activation on the depression phenome (see Figure 1 how these factors were constructed). This model examines the effects of adverse childhood experiences (ACE) and the recurrence of illness (ROI) on the immune variables. ACE was conceptualized as the first factor extracted from mental neglect (MeNeglect), mental trauma (MeTrauma), and physical trauma (PhTrauma). ROI was conceptualized as the first factor extracted from lifetime (LT) suicidal ideation (SI) and attempts (SA), and number of depressive episodes (#episodes). The results of complete PLS analysis are shown as pathway coefficients (p-values) in the inner model, and loadings (p values) in the outer model; the figures in the blue circles indicate explained variance.

### Construction of the final PLS model

**Figure 6** shows the most adequate model in which we built two different pathway-phenotypes, namely a first LV extracted from CD20+CB2+, LT_SA, and ACEsum (labeled: ACE-SB-B cell pathway phenotype); and a second LV extracted from mental neglect, ROI, and the same five T cell activation markers as shown in Figures 1 and 5 (labelled: ACE-ROI-T cell pathway phenotype). Both the ACE-SB-B cell and ACE-ROI-T cell pathway phenotypes showed adequate psychometric properties, including AVE (0.666 and 0.595, respectively), Cronbach’s alpha (0.756 and 0.921), rho a (0.826 and 0.934, respectively), and rho c (0.855 and 0.926, respectively). With an SRMR of 0.080, the model fit is sufficient. PLSpredict shows that all Q2 predict values are > 0.426, and CVPAT shows that the PLS versus indicator average is highly significant (t=4.03, p<0.001). Complete PLS analysis performed on 5,000 bootstraps shows that 71.4% of the variance in the current phenome may be explained by the cumulative effects of Treg, and the ACE-SB-B cell and ACE-ROI-T cell pathway phenotypes.

**Figure 6.**
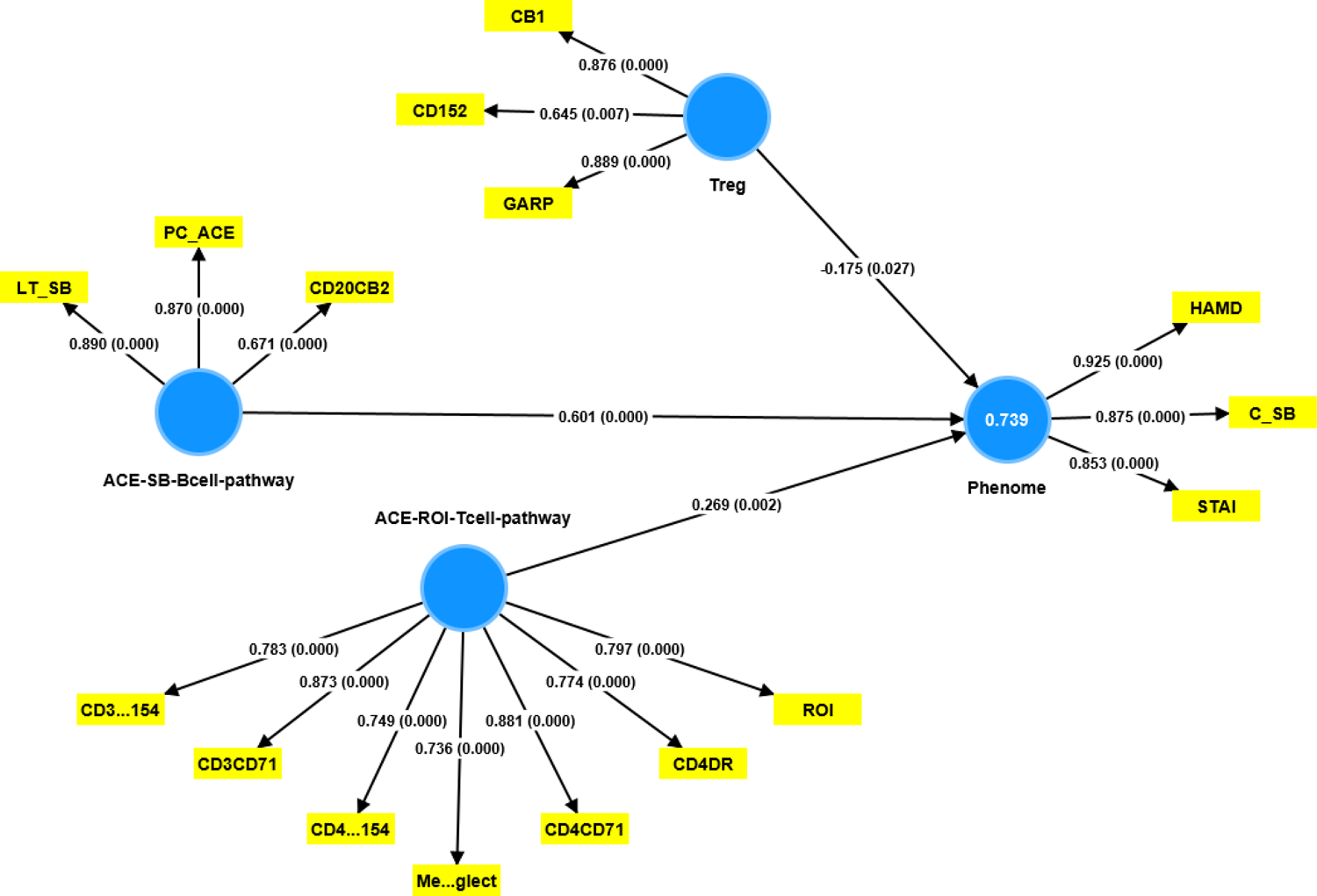
Results of Partial Least Squares (PLS) Analysis. This PLS Analysis shows the most adequate model in which we built two different pathway-phenotypes: a) ACE-SB-B cell pathway phenotype, i.e. a factor extracted from lifetime suicidal behaviors (LT_SB), CD20+CB2+ B cell%, and PC_ACE (a principal component computed using the scores of mental trauma, physical trauma, mental neglect, domestic violence, a family history of mental disease, and the loss of a parent); and b) ACE-ROI-T cell pathway phenotype, i.e., a factor extracted from mental neglect (MeNeglect), ROI and T cell activation markers (see Figure 1 for specifications). The two pathway phenotypes combined with T regulatory cells (Treg; see Figures 1 and 5 for specifications) explain 73.9% of the variance in the phenome of depression (see Figures 1 and 5 for specifications). The results of complete PLS analysis are shown as pathway coefficients (p-values) in the inner model, and loadings (p values) in the outer model.

We were also able to extract one validated LV from LT_SB, number of episodes, HAMA, STAI, Current_SB, CD3+CD71+, CD3+CD40L+, CD4+CD71+ and CD4+CD40L+ (AVE: 0.583; Cronbach’s alpha: 0.912; rho a: 0.928; and rho c: 0.926). We found that 48.8% of the variance in this LV was explained by the ACEs LV.

## Discussion

### T and B cell activation in MDMD

The first major finding of this study is that there are major differences in T cell activation profiles, and number of CD20+CB2+ cells between MDMD, SDMD, and controls. Thus, T cell activation markers (CD71, CD40L and HLA-DR bearing CD3+ and CD4+ cells, and CD8+CD40L+ T cells) and CD20+CB2+ B cells were increased in MDMD versus controls and/or SDMD. Previous findings revealed that MDD is qualitatively distinct from controls as indicated by increased numbers or expression of T cell activation markers (Ig+, CD25+, and HLA-DR+ on CD4+ T cells) and increased numbers of CD19+, CD21+ and HLA-DR bearing B cells as compared with controls [6–8]. These alterations yielded significant accuracy to externally validate the diagnosis of MDD. Thus, 64% of patients with MDD had increased expression of CD7+CD25+ and CD2+HLADR+ cells, with a specificity of 91% [8]. Also, using the various B cell markers showed that 63% of the depressed patients were correctly classified with a specificity of 95% [39]. It should be stressed that the elevated levels of soluble transferrin receptor (sTfR or sCD71) and soluble interleukin-2 receptor (or CD25+) in the serum of MDD and BD patients [1] agree with the current findings. Recently, we detected that first-episode MDMD is accompanied by increased levels of IL-16, which is a CD4 co-receptor ligand that synergizes with IL-2 to induce T cell proliferation [21]. The findings of this study are consistent with the established research on the activation of the IRS in MDD as reported by Maes and Carvalho [3] and in MDMD as reported by Maes et al. [22].

CB2 plays a pivotal role in T-independent humoral immunity and B cell-associated immunity [40]. The upregulation of B cells through CB2 has been observed to stimulate the production of natural IgM antibodies that are directed towards self-antigens [40–42], another hallmark of MDD [43]. As previously discussed, [20] augmented levels of CD20+CB2+ B cells could potentially play a role in the development of autoimmunity and pro-inflammatory responses. Overall, the results indicate that severe depression is characterized by T cell activation and CD20+CB2+ cell proliferation.

### Immune tolerance breakdown in SDMD

The second major finding of our study is that during the acute phase of the SDMD subtype, there is a disruption in peripheral immune tolerance, as indicated by a reduction in Treg cell count when compared to the control group. Furthermore, a distinct reduction in FoxP3+CB1+ cells was observed in the SDMD group as compared to both the control and MDMD groups, while a decrease in FoxP+GARP+ was observed in the SDMD group as compared to the control group. The present study builds upon prior research indicating that individuals diagnosed with MDD exhibit diminished levels of Tregs [16]. Furthermore, Mohd Ashari et al. [17] reported that MDD patients who received antidepressant treatment exhibited a higher count of CD4+CD25+FOXP3+Treg cells compared to those who did not receive treatment. There is a positive correlation between the depletion of CD4+CD25+ Treg cells and the manifestation of depressive-like behaviors in mice [18].

The transcription factor FoxP3 plays a crucial role in the regulation of Treg activities through the production of CIRS cytokines, including IL-10 and TGF-β1, depletion of growth factors, and the expression of the co-inhibitory CTLA-4 molecule. This has been documented in previous studies, such as the one conducted by Lu et al. [44]. Treg cells may express CTLA-4 (CD152), GARP, or CB1, which function as negative immune checkpoints to modulate immune responses [19,20,45]. Workman et al. [46] showed that CLTA-4 can effectively reduce the proliferation and activation of T cells by competing with CD28. GARP regulates the release of TGF-β1, a major CIRS cytokine [47]. Tregs have been found to express CB1, which has been shown to attenuate the production of pro-inflammatory cytokines, including IL-2 and IL-12, and trigger apoptosis in immune cells [48].

Besides depletion of Tregs, SDMD is accompanied by a depletion of CD3+CB2+ T cells as compared to MDMD and the control group, whilst CD4+CB2+ T cells were found to be lower in SDMD in comparison to MDMD. The maintenance of immune homeostasis can be achieved through the ligation of CB2 or the upregulation of CB2 expression on T cells [49–53]. The observed impacts involve both anti-inflammatory and anti-proliferative effects, which are achieved through the reduction of CD4+ cell proliferation, the production of Th-1 cytokines (IFN-γ and IL-2), T effector activities and T cell functions, Th-17 activation, macrophage and chemotaxis, and the induction of Th-2 polarization. As such, the decreased presence of CB2-bearing T cells in SDMD may potentially aggravate the disruption of immune tolerance because of Treg depletion.

A noteworthy finding of the present investigation is that a considerable proportion (approximately 40%) of the variability in the depression phenotype, encompassing depression, anxiety, and suicidal behaviors, can be accounted for by the collective impact of T cell activation, augmented CD20CB2+ cell counts, and diminished Treg cell counts. Thus, indicators of T cell and B cell activation in conjunction with immune tolerance breakdown are associated with a more severe phenome, including suicidal behaviors.

### MDMD versus SDMD

Previous research showed that MDMD is characterized by elevated pro-inflammatory cytokine and growth factor profiles, as well as increased IRS-associated neurotoxicity, whereas these features are not observed in SDMD [22]. The same authors [23,24,27,54,55] have reported that MDMD exhibits several distinct characteristics that are not present in SDMD. These include heightened IgM autoimmune responses to self-antigens, which suggest increased B1 activity, as well as elevated levels of oxidative stress, reduced lipid-associated antioxidant defenses, increased gut dysbiosis and translocation of Gram-negative bacteria, and a lowered neurotrophic protection. The MDMD subgroup is not only the biologically disordered depression phenotype, but also presents a clinically more severe phenotype, exhibiting more severe depression, anxiety, suicidal behavior, lower quality of life scores, and a greater ROI compared to the SDMD subgroup.

It is noteworthy that the current study indicates that SDMD exhibits distinctive attributes, which differentiate this subset from both MDMD and control groups, including lowered Treg and CD3+CB2+ and CD4CB2+ cells. Importantly, these results suggest that SDMD is associated with decreased immune tolerance, which may lead to relatively increased IRS activation. Recent findings indicate that the SDMD subgroup is distinguished by a distinct reduction in the CIRS cytokine profile, which encompasses a decline in IL-4 levels (Maes et al., 2023, TBS). Therefore, it can be inferred that SDMD is not solely a minor manifestation of MDMD, but rather possesses distinct characteristics unique to itself.

### Effects of ROI on T and B cell activation and immune tolerance

The third major finding of the present study is that ROI (and the number of depressive episodes and suicidal behaviors) is strongly associated with CD20+CB2+, T cell activation (CD3+CD71+, CD4+CD71+, CD3+40L+, CD4+CD40L+, and CD4+HLADR+), while the number of episodes is inversely associated with Treg counts. Overall ROI has a very strong impact on the combined T + B cell activation, and T + B / Treg ratio indices, with an estimated effect size of around 37% explained variance. Thus, with increasing ROI, the degree of T and B cell activation increases gradually, indicating that the ROI is accompanied by an increasing dysbalance in T and B cell activation versus Treg protection.

These results extend previous papers on the associations between ROI and immune and immune-related biomarkers of depression. Early research showed that serum levels of IRS, such as neopterin, IL-6, and CIRS (IL-10, sIL-1RA) biomarkers, are associated with the number of depressive and manic episodes in MDD and MDE [56–60]. Recently, we detected that the ROI is associated with activation of different immune profiles, including M1 macrophage, Th1, Th2, Th17, IRs, CIRS, T cell growth, growth factor, and neurotoxicity profiles [25]. Nevertheless, the strongest impact of ROI on these profiles was established for T cell growth cytokines, including IL-4, IL-9, IL-12, IL-15, and GM-CSF, which as such extends the findings of the current study. These findings indicate that with increasing ROI, the immune system of depressed patients becomes more sensitized. The phenomenon of time-dependent sensitization is well-established in response to exposure to stressors, whether they are of psychological or organic nature [61–63]. This immune sensitization is contingent upon a re-stressor event and is observed to augment progressively over time. The kindling theory of affective disorders, as posited by Post [64], suggests that with each recurring episode, sensitization increases. Using the precision medicine approach, Maes et al. [25] showed that one factor could be extracted from ROI, the depression phenome, and the T cell growth, IRS, and neurotoxicity profiles, thereby generating an ROI-immune-phenome pathway phenotype. Likewise, pathway phenotypes could also be constructed by extracting a latent vector from ROI and lowered antioxidant defenses and oxidative damage biomarkers, and from ROI, the phenome, and a gut microbiota enterotype [23-25,27,54,55]. All in all, the results of our study indicate that the phenomenon of “affective kindling” is attributable to anomalies in T cell activation, IRS activation, redox-related pathways, and a gut microbiome dysbalance.

### Effects of ACEs on ROI, T and B cell activation, and immune tolerance

The study’s fourth significant discovery indicates a positive and significant correlation between ACEs and ROI and various immune cell counts, including CD20+CB2+ B, and CD3+CB2+, CD4+CB2+, CD3+CD40L+, and CD4+CD40L+ T cells. Conversely, there was an inverse relationship between ACE and Treg cell counts. The findings indicate that ACEs had a significant impact, accounting for as much as 22.8% of the variability observed in the T + B / Treg ratio. In prior studies conducted by Maes et al. [25,26,27,54,55], it was demonstrated that an elevation in ACE levels was concomitant with an increase in ROI scores, a reduction in antioxidant defenses, an increase in lipid peroxidation and oxidative damage to proteins, and a microbiota enterotype that indicated the presence of gut dysbiosis. The findings of our study indicate the possibility of creating two separate pathway phenotypes utilizing ACEs, immune, and ROI data. These phenotypes are as follows: a) ACEs – lifetime suicidal behaviors - CD20+CB2+% pathway phenotype, and b) ACEs – ROI – T cell activation pathway.

These discoveries demonstrate a significant correlation between ACEs, ROI, and activated T and B cell pathways, which give rise to distinctive ACEs-ROI-immune pathway phenotypes. Furthermore, our research revealed that the combination of both pathways, along with decreased Treg protection, accounted for 71.4% of the variability in the depression phenome. These observations indicate that increased ACEs, ROI, and T and B cell activation are the key components of MDMD, and lowered Treg in SDMD and MDD.

### Limitations

The study could have been enhanced by the inclusion of measurements of oxidative stress and gut microbiota biomarkers as well as the enumeration of other immune cell types, such as natural killer cells, macrophages, and dendritic cells. Subsequent studies ought to examine the associations between the immune cell subtypes assessed here and depression in the partial remission and remission phases. One could posit that the size of the sample is relatively limited, thereby reducing the accuracy of the parameter estimates. However, it is noteworthy that the a priori minimum sample size estimation was established through the utilization of power analysis. Additionally, a post-hoc power analysis was conducted on the primary analysis, which involved the regression of the phenome on immune cell populations (Figure 1). The results indicate that the power obtained from the current primary analysis is 0.998.

## Conclusions

The acute phase of SDMD is typified by a disruption of immune tolerance resulting from diminished levels of regulatory T cells (Treg), CD3+CB2+ and CD4+CB2+ T cells. Meanwhile, MDMD is characterized by T cell activation associated with CDL40+ and CD71+, as well as heightened levels of CD20+CB2+ B cells. In general, the impairment of immune homeostasis in depression may play a role in the activation of the IRS that is commonly observed in this condition. Elevated levels of IL-16 may lead to heightened T cell activation, subsequently inducing the activation of CD8+ cells (CD8+CD40L+) and promoting the acquisition of additional CB2 receptors by B cells. The latter process may result in excessive production of IgM antibodies and trigger autoimmune and inflammatory responses [19,20]. Heightened ROI is associated with the activation of T effector (CD4+), T cytotoxic (CD8+), and CD20+ B cell subtypes, as well as the reduction of Treg protection, which together cause IRS activation and increased neurotoxicity [19,20]. This indicates that the ROI is linked to a sensitized disbalance among detrimental lymphocyte phenotypes (activated effector and cytotoxic T cells and CB2-bearing B lymphocytes) versus negative immunoregulatory phenotypes (Treg). Consequently, autoimmune processes are probably a major factor in the recurrence of depressive episodes and suicidal behaviors. Furthermore, ACEs may play a pivotal role in these detrimental immune processes that are a defining characteristic of depression by driving immune-associated neurotoxicity.

## Ethical statement

All subjects gave their informed consent for inclusion before they participated in the study. The study was conducted in accordance with the Declaration of Helsinki of 1975, revised in 2013, and the protocol was approved by the Ethics Committee of the Institutional Review Board of the Faculty of Medicine, Chulalongkorn University, Bangkok, Thailand (#528/63).

## Data Availability Statement

The dataset generated during and/or analyzed during the current study will be available from the corresponding author (M.M.) upon reasonable request and once the authors have fully exploited the dataset.

## Author’s contributions

Design of the study: MM and MR. Recruitment of the participants: MR and KJ. Assays: PS. Statistical analyses: MM. Visualization: MM. Editing: MR, KJ, AS, and PS. All authors agreed to publish the final version of the manuscript.

